# The Association of Demographic Factors to Health Policy Subscriptions in Saudi Arabia: insight from National Survey

**DOI:** 10.1101/2025.01.27.25321187

**Authors:** Ahmed Ali Alasiri, Qi Zhang, Mohammed Sahabi

## Abstract

**Background:** Saudi approaches to delivering health services seem not sustainable, and it may limit access to the necessary healthcare services. Therefore, the health system is undergoing several paradigms shifts with the investment of the private sector and insurance. The subscriptions to the health policy are affected by several variables, while demographic indicators of Health policy subscription among Saudi and expatriate populations remain underexplored.

**Aim:** The study aims to test the association of the age, gender and nationality on the subscription of health policy by utilizing national data.

**Method:** The study retrieved secondary data from Saudi Data & AI Authority (SDAIA), collected between 2021 -2022 with a nationality sample that cover all Saudi regions (n=14.780). The project utilized descriptive and inferential statistical tests to analyze the data and understand the relationship between the demographic variables and how they affect the subscription to health insurance.

**Result:** While age, gender, and nationality have impacted the subscription of insurance to varying degrees, however, these factors revealed a small effect on the policy subscription, indicating that other social-economic variables may have a stronger prediction of the subscription rate.

## Introduction and Background

Delivering healthcare services for the population is a complex endeavor. The health services affected directly or indirectly way by countless factors. In theory and the ideal approach of utilizing health service, need should be the main and first health determine of health utilization among patients (1). The government in developed and developing countries strived to established and advanced strategies to face these challenges. Kingdom of Saudi Arabia (KSA) initiated health transformation for its health system to ensure sustainability and to address current challenges (2,3)

Also, countries have several approaches to deliver health services for its nations, KSA is one of the few countries that provide and deliver care services for citizen and expatiate for free that include primary, secondary and tertiary health care services (4) .However, this approached may hinder the improvement of the quality and it stands as challenge for long -term sustainability.

When examine the population characteristics in KSA, a major challenge emerges with percentage of expatiate who work in the private and government sector. They represent about 41.7 % of the total population, and private scoter did not do its share by providing health services for its employees while the demand of services increased over years (5,6).

In the last twenty years, the health insurance legislation and provision have founded to ease the burden on public sector. Initially, it is started with an attempt to cover employees in private sector. The council of the Cooperative Health Insurance established law in 1999 to provide a mandatory health insurance for people who work in private sector that include citizen or non-citizen (7). The insurance provision gradually improves the number of the insured to reach 11 million (8,9). Comprehensively, Health care system is undergoing a substantial change to improve the access to health care and development the standard of health to align with international benchmark (2,8–10).

While health insurance established in the country and it is mandatory for employees in the private sector, with increases of the investment of the private insource to reach 17 companies, however the demographics characteristics of individual and how that associate to the subscription to the health insurance have limited discovered in the literature (8). Therefore, this project focused on studying factor that might affect inequality to the health services though health insurance. Further, this insight could lead to tailoring intervention to improve health insurance literacy among population. This research is driven by the need to explore the relationship between the core demographic variables (age, gender and nationality) on the subscription to the health insurance, within the current health care transformation.

## Methods

### Study setting

The study conducted in one of the largest countries of the middle east, Kingdon of Saudi Arabia; it is in Western Asia, and it covered about 2.15 million km of the Arabian Peninsula (12). It has population 32.175.224 with the average age 29 years. The percentage of Citizen represent 58.4% and to non-citizen 41.6% (13,14). KSA is one of the top twenty economy in the world (15), and it is categorized a high-income and high human development index economy based on the World bank (16)

## Database

The data was retrieved from Saudi Data & AI Authority (SDAIA). it is a secondary data, and it is available for public (17,18). The data includes 140.792 participants that covered 13 administration area in the KSA. The data contains three independent variables and one outcome. The Independent variables include age, gender, and nationality and there is only one dependent variable which is the number of the subscription to the health policies. The data collected in 2021, and it has been updated on the second quarter of 2022 year.

## Data management

The data retrieved as excel format, it was on English language. The total number of samples included in the study is 140.729. Microsoft excel utilized to code the data as following .Age classified to 14 gropes 1- (0-5) 2- (6-10), 3- (11-15) ,4- (16-20) 5- (21-25) ,6- (26-30),7- (31-35),8- (36-40),9- (41-45),10- (46-50),11- (51-55),12- (56-60), 13- (61-65) 14-More than 65. Gender coded as 1-Male ,2-Female. Also, Nationality categorized as Saudi -1 and non-Saudi -2. There is no missing data.

### Statistical Analysis

Descriptive of the data to analysis the demographics of the people include characteristics, frequencies and percentages. One-way ANOVA utilized to examine the effect of age gropes on the subscription of the health policy. Also, Independent T-test was conducted to test the mean differences between gender on total subscription. Likewise, the independent t-test was performed to compare the mean between citizen and non-citizen on policy subscription. Additionally, multiple liner regression was conducted to investigate the relationship between the independent variable (age, gender and nationality) and the dependent variable (total number of subscripting to health policy). The significant level was set < 0.5, and the CI 95%. The project utilized SPSS version 28 (19).

## Result

There are 14 gropes of ages, and the people age range from 0 to more than 65 years old. The majority of responses are between 21-45 years old, and it represent 46.4 %, while people from 0-5 and more than 65 year and older represent only 9.7%. people range between 6-20 years old show about 19.1 %. Also, age grope between 46-65 years is 24.7 %. Male and female have similar percentage while Male account 50.7% and female 49.3%. Non-Saudi have more subscription than Saud 57%-42% respectively (Table 1).

**Table 1:** Demographics characteristics of study groups.

**Demographics characteristics of study groups (Table 1)**
| Characteristics | Frequency | Percentage | Mean | *P*-value |
| --- | --- | --- | --- | --- |
| Age (years) |  |  |  |  |
| 1- (0-5) | 8294 | 5.9 % |  |  |
| 2- (6-10) | 8487 | 6.0 % |  |  |
| 3- (11-15) | 8123 | 5.8 % |  |  |
| 4- (16-20) | 10310 | 7.3 % |  |  |
| 5- (21-25) | 13153 | 9.3 % |  |  |
| 6- (26-30) | 13252 | 9.4 % |  |  |
| 7- (31-35) | 13446 | 9.6 % |  |  |
| 8- (36-40) | 13290 | 9.4 % |  |  |
| 9- (41-45) | 12304 | 8.7 % |  |  |
| 10- (46-50) | 10716 | 7.6 % |  |  |
| 11- (51-55) | 9605 | 6.8 % |  |  |
| 12- (56-60) | 8237 | 5.9 % |  |  |
| 13- (61-65) | 6207 | 4.4 % |  |  |
| 14- More than 65 | 5368 | 3.8 % |  |  |
|  |  |  | 7.21 | <.001 |
| Gender |  |  |  |  |
| **Male** | 71404 | 50.7 % |  |  |
| **Female** | 69388 | 49.3 % |  |  |
|  |  |  | 1.49 | <.001 |
| Nationality |  |  |  |  |
| **Saudi** | 59659 | 42.4 % |  |  |
| **Non-Saudi** | 81133 | 57.6 % |  |  |
|  |  |  | 1.58 | <.001 |

A one -way ANOVA result found that the age gropes have statistically significant differences of the subscription for health policies, F (13, 140778) = 16.03, p < .001 however the effect size between the 14 gropes of age were small with eta-squared (η^2^) = .001. in another word, the impact of age to subscription of health policy is not notable and it represent only 0.1% of variance of health policy subscription. Independent T-Test conducted to test the difference in subscription to health policy between Saudi and non-Saudi population. The result found that there is a statistically significant differences between two gropes, t (137,393) = -5.95, p < .001 with the mean difference between gropes was -31.83, 95% CI [-42.31, -21.35]. However, the actual impact of nationality as illustrated by effect size was very small Cohen’s d = -0.031. Also, the independent t-test found that there was a statistically significant of the gender in the policy subscription t (100,645) = 15.07, p < .001. Male had higher mean than female of subscription, and the difference between mean was 80.71 (95% CI [70.22, 91.21]).However ,the effect size was small, Cohen’s d, 0.080 .which indicate that there was only 0.8% effect of the age on the subscription to the health policy. Multiple Regression was conducted to examine the ability of the age, gender and nationality to predict the number of policy subscription.

The model summary found that the overall fit of regression was low R = .044, while the R^2^=.002 which showed that only 0.2% of the variance explained by the age, gender and nationality factors. ANOVA table discovered that overall model was statistically significant F (3,140788) =90.460, p<.001. The coefficient table showed that age was significantly statistic -3.636 (p < .001), and it showed that one additional year in age for each grope, there is a decrease of subscription to health policy by 3.636. The coefficient for nationality was statistically significant 28.715 (p < .001), and it indicate that being non-Saudi increase the subscription of health policy by 28.715. Also, the gender was statistically significant -79.771 (p < .001), which indicating that being female decrease the subscription by 79.7

**Table 2:**
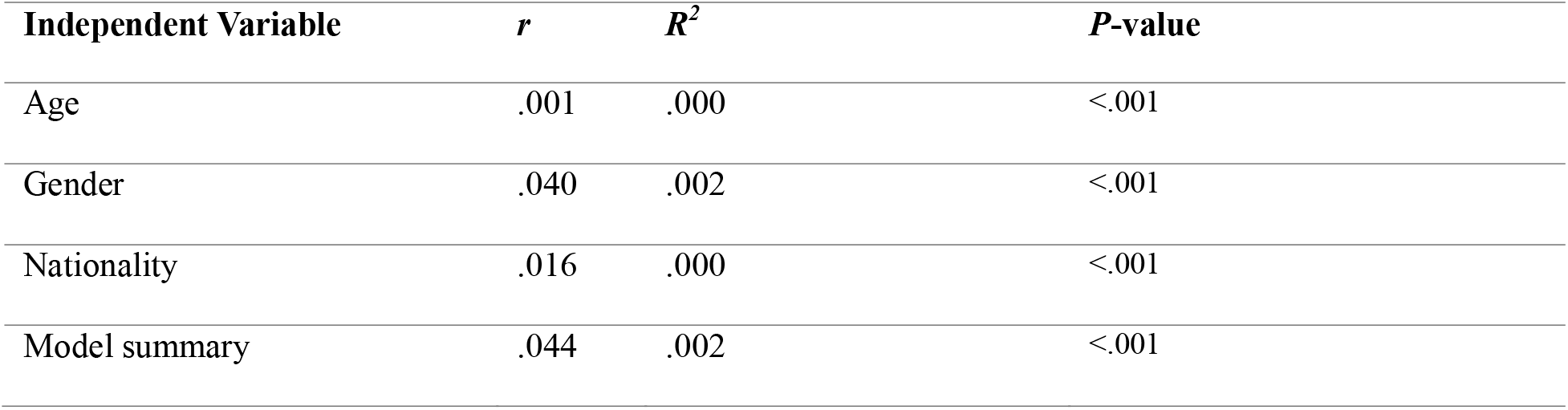
The coefficient between demographics factors and the number of subscriptions to health policy.

## Discussion

This project aimed to study the association of the demographics variables (age, gender and nationality) to health policy subscriptions. The test of variances include One way ANOVA and Independent t-test found that the age gropes, gender and nationality have significant impact on the health subscriptions however the effect size of them were very small. Similarly, the result of the multi regression found that the age, gender and nationality predicted the number of subscriptions however the overall model showed small proportion of variance in the subscription to health policy.

### Age as a Determinant

The trends from various studies indicate that age is a major factor in health insurance subscriptions. According to Al-Hanawi stated that an older person is likely to subscribe to health insurance in Saudi Arabia because of the increased morbidity rate and increased prevalence of chronic illnesses (20) . while present study found similar pattern with effect of the age factor, however the number of subscriptions decreased when participants get older.

Aljehani et al. observed that there has been a high utilization of health insurance by elderly Saudi citizens, especially those suffering from chronic diseases such as diabetes. (21). Altogether, the analyses of researchers from other countries are not very different from the indicated patterns. In Germany, where insurance is mandatory, especially for older people, health insurance entails broad coverage as older people use health facilities more than younger people (22). On the other hand, accession to older generations is less frequent. It is scheduled later in life, given the relatively low sensitivity to potential health threats in the United States (23).

### Gender Disparities

Sexual differences in receiving health insurance are well known. Alatinga demonstrated that gender bias in health insurance by showing that cultural and economic barriers make it hard for women to access health insurance programs (24). Similarly, current study observed that male has more subscription to insurance regardless their nationality. Research from high-income countries documented more complex patterns. For example, even in a country with insurance for everyone, like Canada, women have equal chances of getting insurance but have unequal chances of getting specialized insurance for maternal and reproductive health services. Such comparisons show the need to focus on extending the programs to remove the system barriers, hence providing Gender equality in access to health policies (25).

### Nationality and Expatriate Consideration

Due to the significant number of foreigners in the country, nationality is one of the most important factors in the KSA now. It has come to be realized that all Saudi nationals are free to access government-funded health care. At the same time, the expatriates depend on health insurance plans sponsored by their employers, which are comparatively limited. Alkhamis explained that while expatriate workers in Saudi Arabia lack coverage or get inadequate coverage, the situation differs from that of countries such as the UAE (26). The UAE requires employer-provided health insurance for expatriate, meaning higher coverage (27). Similarly, research conducted in Singapore explains how government subsidies that increase expatriate insurance plans help non-citizens (28). These examples show that expatriates’ concerns, which are attended to by inclusive policies, can considerably decrease the gaps, which Saudi Arabia can apply to improve the health insurance system as per Vision 2030.

## Implications and Future Directions

The study underscores the importance of the government further looking into the issue of demographic differences in health policy subscriptions. There is a need to develop good strategies like providing subsidies to serve such high-risk categories and introducing measures to cover expatriates comprehensively through health plans. This is the perfect time for Health Vision 2030 to steer the health policies toward principles of inclusion and equity. Further studies should determine the effectiveness of innovative digital health strategies and the roles of PPPs in closing these demographic disparities.

## Conclusion

The independent variables of study include Age, gender and nationality have influenced the subscription to the health insurance by varying extent. one year extra of participant age related to reduce number of subscriptions. Expatriates have more effect than Saudi citizen on the outcome. Male gender has more subscription to female with low effect size for all of them. While KSA focused on affordability and elevate access to high quality of health services ,however further scientific researches needed to discover other factors that may significantly influence subscription to health policy .

## Data Availability

All data produced in the present study are available upon reasonable request to the authors

